# Deep Learning Driven Field Dose Prediction for Head and Neck Cancer Treated with Spot Scanning Proton Therapy

**DOI:** 10.64898/2025.12.08.25341831

**Authors:** Brandon Reber, Satomi Shiraishi, Andrew Y. K. Foong, David Routman, Jing Qian

**Author notes:** **Correspondence** Brandon Reber.

## Abstract

**Purpose:** Accurate dose prediction is essential for automating radiotherapy planning. In spot scanning proton therapy (SSPT), dose evaluation is required at both the plan and field level. Evaluating individual treatment fields is critical to ensuring optimal beam angles are chosen to ensure target coverage and maximum organ-at-risk (OAR) sparing. Currently, however, no knowledge-based tools exist for predicting field-level doses for head and neck cancer (HNC) treated with SSPT. In this work, we aim to develop the first deep learning–based dose prediction model capable of field-level dose prediction for HNC treated with SSPT.

**Methods:** A cohort of 62 HNC patients treated with SSPT was compiled for model development and evaluation. Collected patient data included treatment planning CTs, OAR masks, signed distance maps (SDMs), generated beam masks, and dose distributions. An encoder-decoder architecture enhanced with a cross-attention transformer bottleneck was used as the field prediction model. Comparison and ablation studies evaluated the model’s performance and determined the benefits of individual model components. Evaluation imaging metrics included mean absolute error, structural similarity index measure, and peak signal-to-noise ratio. Clinical performance was evaluated using dose-volume histogram metrics.

**Results:** The best performing model from the ablation study was the full model using OAR masks, SDMs, generated beam masks and four-field dose prediction. The model outperformed the Distance Guided Dose Prediction (DGDP) and DeepLabV3 comparison models. The DGDP and DeepLabV3 comparison models had a mean validation set MAE performance of 1.268 Gy and 1.325 Gy, respectively, compared to our model’s mean validation set MAE performance of 0.949 Gy. The model’s final mean test set performance was MAE 1.024 Gy, SSIM 0.913, and PSNR 28.495 dB.

**Conclusions:** We developed a cross-attention transformer–enhanced deep learning model that accurately predicts per-field dose for HNC treated with SSPT, demonstrating superior performance over state-of-the-art models limited to plan-level dose prediction.

## Introduction

Proton therapy is a radiation treatment modality that is increasingly used in the United States. From 2012-2021, the number of head and neck cancer (HNC) patients treated with proton radiotherapy in the United States grew by approximately 600%^1^. An increasing number of patients treated with proton therapy highlights a growing need for tools to assist in the development of treatment plans. Automated tools such as deep learning models have the potential to not only reduce the time to develop radiation therapy plans but improve plan quality as well^2^.

Proton-based radiation therapy differs from photon-based radiation therapy in both the physics of dose deposition and in treatment planning considerations. Photons deposit energy in a broad dose fall-off compared to protons that deposit a large amount of dose in a relatively short distance (Bragg peak)^3^. For photon plans, setup uncertainty is typically incorporated into treatment planning by using a planning target volume (PTV)^4^. Range and setup uncertainties in proton plans lead to PTV alternatives for targeting uncertainties such as robust optimization^5^. Volumetric modulated arc therapy (VMAT) is a common photon treatment technique to deliver highly conformal treatment using one or more delivery arcs. In comparison, many proton techniques such as passive scattering and spot scanning proton therapy (SSPT) are delivered using a series of fixed beam angles^6^.

Deep learning has been applied to a variety of HNC cancer related tasks such as tumor segmentation, late toxicity prediction, modality decision making, and HNC tumor molecular status characterization^7 8 9 10^. In particular, deep learning-based radiation dose prediction methods have largely replaced dose prediction methods based on earlier machine learning techniques. Deep learning has several advantages over traditional machine learning such as allowing full images as model input and by automating feature extraction from medical imaging^11 12^.

Previous work in deep learning-based HNC dose prediction has mainly focused on photon-based treatments^13 14 15 16 17 18^. Many methods use the open source OpenKBP dataset of HNC patients treated with photon radiation therapy for model input rather than institution specific cohorts^19^. Several models use variations of the standard U-Net architecture for dose prediction^13 15 16 17^. Some studies employ other deep learning architectures for HNC photon dose prediction including generative adversarial networks and transformers^14 18^. Fewer studies have examined using deep learning for proton dose prediction. Studies that do exist use standard variants of U-Net architectures without the potential benefits of newer, advanced architectures such as transformers^20 21 22 23 9^.

Proton therapy has distinct features compared to photon therapy that presents unique considerations when developing deep learning dose prediction models. Unlike photon VMAT, most proton therapy treatments employ a restricted set of beam angles and produce much sharper dose gradients at both beam edges and distal falloffs. These characteristics result in inherently discontinuous dose distributions, which pose distinct challenges for deep learning–based dose prediction. In addition, each proton field contributes uniquely to overall plan robustness, biological effectiveness, and organ-at-risk sparing. Therefore, evaluation of plan quality and dose distribution must extend beyond the conventional plan-level assessment to include detailed field-level analysis. This is further underscored by the fact that machine delivery parameters for proton therapy are defined at the field level, not at the plan level, making accurate field dose estimation a prerequisite for translating dose prediction into clinically deliverable proton plans. Despite this critical need, existing knowledge-based and deep learning models remain confined to plan-level dose prediction, neglecting the essential role of individual field contributions. To date, no model has been developed to predict head and neck cancer proton dose distributions at the field level.

In this study, we present an innovative dose prediction model developed using an institutional HNC cohort treated with SSPT and evaluated using both imaging metrics and clinical metrics. This work advances beyond previous studies in several key aspects. First, the model predicts dose distributions not only at the plan level but also at the per-field level, delivering critical field-specific information absent from prior approaches. Second, unlike earlier proton dose prediction architectures, our design incorporates a modern encoder–decoder framework enhanced with a cross-attention transformer bottleneck, enabling superior representational capacity and performance. Third, we systematically evaluate the contribution of individual input channels through comprehensive ablation studies, providing quantitative evidence to guide the optimal construction of model inputs.

## Methods and Materials

### Patient cohort

A retrospective cohort of patients treated at Mayo Clinic Rochester from 2015-2024 was identified for model construction and evaluation. The patients had bilateral oropharynx cancer treated with definitive radiotherapy using a prescription dose of 70 Gy delivered in 35 fractions. Treatments were planned with CTV high receiving 70 Gy, CTV intermediate receiving 63 Gy, and CTV low receiving 56 Gy.

The patients were screened to ensure that four proton fields were used for treatment. Most HNC proton treatments at our clinic use three to five treatment fields, with the majority of patients receiving treatments consisting of four or three fields. Treatments with different numbers of fields have different field weightings to deliver the required total prescription dose. To ensure that no model prediction errors were associated with incorrect field weightings, the four field treatment geometry was exclusively used in our model. The four treatment fields are oriented at the left anterior oblique, right anterior oblique, anterior, and posterior positions. Figure 1 depicts an example of a typical four-field proton beam arrangement for the included patients.

**Figure 1:**
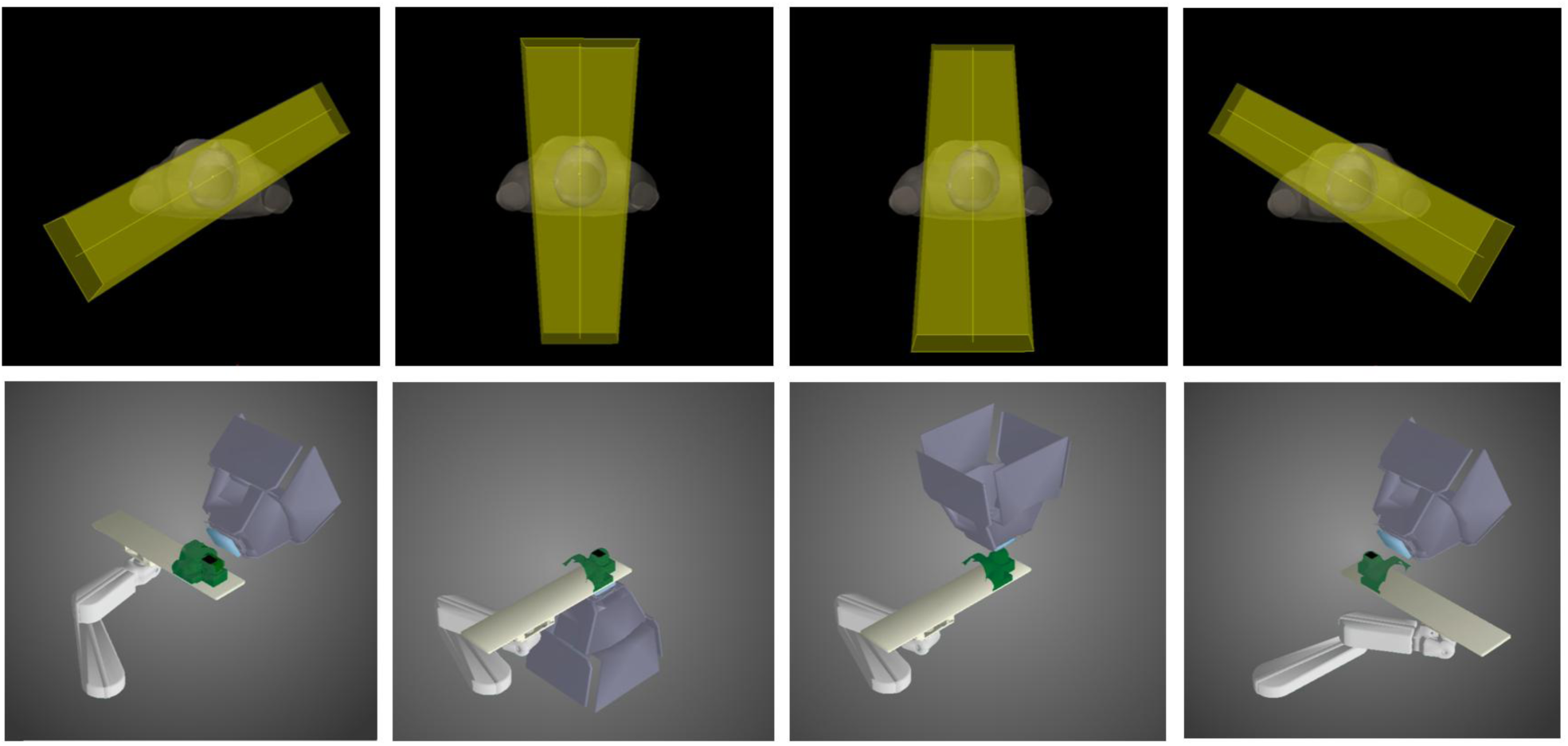
Examples of four field beam geometry for proton head and neck cancer radiation therapy. Treatments typically consist of a right anterior oblique (column 1), posterior (column 2), anterior (column 3), and left anterior oblique (column 4) fields. Each column corresponds two views of the beam geometry. The green ROI on the patient couch in the bottom row is an example of an external patient contour.

For each patient, the ground truth dose was generated using an in-house Monte Carlo algorithm^24 25^. Patient treatment planning CTs were acquired along with region of interest (ROI) binary masks. ROI binary masks included CTV high, CTV intermediate, CTV low, brainstem, esophagus, larynx, parotids, mandible, spinal cord, and external masks. External masks are masks of the body with a 1 cm expansion. Patients with missing OAR masks had masks of empty arrays created. For all OAR masks, signed distance maps (SDMs) were created. SDMs give each voxel in a ROI mask the value that is the closest Euclidean distance to the contour boundary. For our implementation, SDM voxel values are positive for values within a mask and negative for values outside a mask. Blank ROI masks received a blank SDM.

### Field Mask Generation

Unlike ROIs, treatment field dose maps do not exist when the model is used at inference time, and therefore, an alternative to creating binary masks from ground truth proton fields must be used. Prior to treatment planning, for our clinical setup, each field’s geometry is specified by a gantry angle and a treatment couch angle. The gantry angle describes the rotational position of the gantry around the treatment couch, and the treatment couch angle describes the rotation of the patient along an axis perpendicular to the rotation angle of the gantry. These two angles were used to create a binary mask for each field similar to the ground truth binary field mask.

To generate the field binary masks, first, a combined CTV mask was generated from the union of the patient’s CTV low, CTV intermediate, and CTV high masks. After translating the imaging isocenter to the treatment isocenter, the combined mask was then rotated by the corresponding gantry angle followed by the treatment couch angle. After rotation, the array indices corresponding to the largest non-zero mask widths on the x axis and z axis are found. The voxel index along the y axis that corresponds to the largest y axis nonzero voxel value was also found. A padding of 10 indices was added to all selected indices to increase the generated field mask size in all directions around the combined CTV. Next, voxels were filled along the y-axis from the largest nonzero y index back to the zero y index. Voxels are only filled if the x and z voxel indices are within the previously found x and z indices. After the voxels were filled, the mask was then rotated back by the treatment couch angle followed by the gantry angle. The images were then translated back to the original imaging isocenter. An illustrated example of the mask generation process is in the supplementary material (Figure S1). In addition to treatment field masks, SDMs of the field masks were created. SDM creation for the field masks followed the same procedure as other ROI masks.

### Image Preprocessing

All images were resampled in the transverse plane to 256x256 voxel dimensions. After resampling, we ensured that images of the same type, such as CT, had the same voxel spacings in all the collected patient images. CT images were clipped to the range of -1000 HU to 3000 HU and then divided by 1000. The ground truth dose images were clipped to 0 Gy and 80 Gy before being divided by 80. Next, to accommodate the model’s 2D input and output, 2D transverse slices were extracted from all images.

### Architecture Description

The dose prediction model used is an alteration of the DoseDiff architecture from Zhang et al^26^. This is a modified U-net architecture enhanced with a cross-attention transformer bottleneck. Three encoder branches were used for the CT, masks, and fused image features. The mask encoder input consisted of concatenated OAR masks, OAR SDMs, treatment field masks, and treatment field SDMs. The fused image feature encoder branch consisted of CT and mask encoder data fused at multiple feature resolutions. Skip connections joined the fused feature encoder with the single decoder branch. The bottleneck consisted of a cross-attention transformer. The cross-attention transformer had the CT branch features as keys, the fused image feature branch as values, and the mask encoder branch as queries. The model has four output channels corresponding to the four individual proton field doses. The total dose for the plan is obtained through summing the four dose output channels. Figure 2 shows a summary of the model architecture. The supplementary information contains additional details about the architecture implementation.

**Figure 2:**
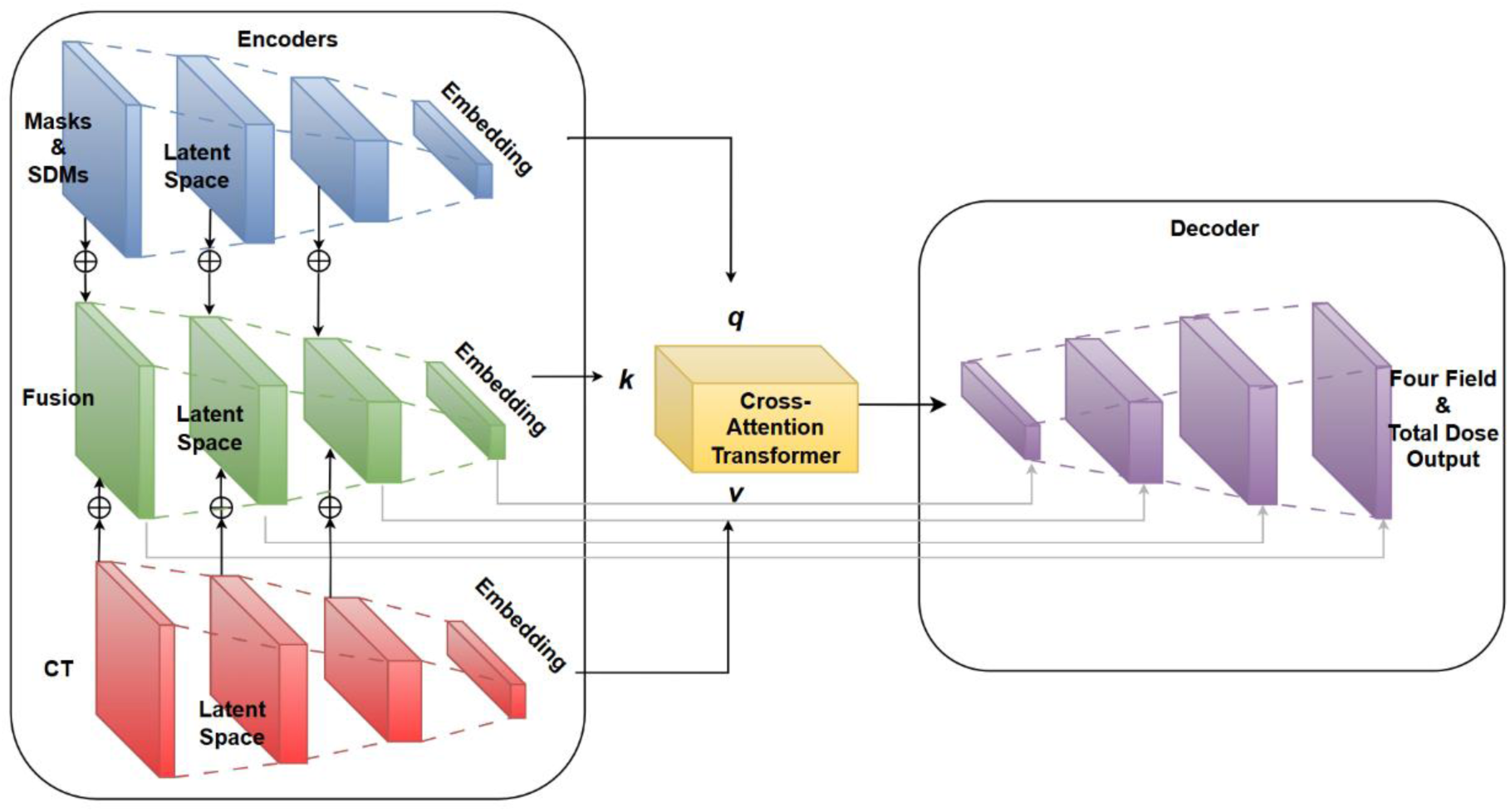
Summary of Model Architecture. The backbone of the model consists of a U-Net model enhanced with various features. There are three branches to the model. One branch uses concatenated ROI masks and signed distance maps (SDMs), one branch uses treatment planning CTs, and one mask uses the fusion of other branch features at multiple resolutions. The bottleneck of the model uses a transformer. The output of the model has four channels corresponding to four treatment fields. The total plan dose is obtained by summing the dose from the four fields together.

### Ablation and Comparison Studies

We compared two established dose prediction networks to our dose prediction algorithm. The two comparison models were models based on the DeepLabV3 network and the distance guided dose prediction (DGDP) network^27 28^. The two models used model implementations and training details as close as possible to their original implementations. For the ablation study, four model components were tested. These model components were the generated masks, the SDMs, the ROIs, and the four field dose prediction.

### Training details

The model was trained using the AdamW optimizer with an initial learning rate of 1e-4 and L2 weight decay. Data augmentations included horizontal and vertical flipping. The mean squared error loss function was used with all four output channels. Models were trained for 500 epochs in total. A step learning rate scheduler was used with the learning rate decreased by a factor of 10 every 125 epochs. Models were trained using four Nvidia A100 GPUs with a batch size of 64 split between the four GPUs. The validation set was used to select the best performing model during training using the mean squared error between the predicted dose and ground truth dose. Descriptions of training for the two comparison models, DeepLabV3 and DGDP, can be found in the supplementary information. For the ablation studies, the same training details were used as the complete model except for the model removing the four-field dose prediction. The model that removed the four-field dose prediction had a single output channel, the total ground truth dose, and the mean squared error loss function was computed between the total predicted dose and total ground truth dose.

### Evaluation metrics

Both standard imaging metrics and clinical performance metrics were used to evaluate models’ dose prediction performance. For imaging metrics, metrics were only evaluated within the external contour. The imaging metrics included were the mean absolute error (MAE), the peak signal-to-noise ratio (PSNR), and the structural similarity index measure (SSIM). Dose-volume histogram (DVH) metrics evaluated the model’s clinical performance. The DVH metrics evaluated were chosen to represent clinically important DVH metrics. The reported values are the difference in the DVH metric between the predicted dose and the ground truth dose. The following DVH metrics were chosen: brain stem (ΔD_max_[Gy], ΔV_30Gy_[%]), spinal cord (ΔD_max_[Gy]), esophagus (ΔD_mean_[Gy], ΔV_55Gy_[%], ΔV_35Gy_[%]), larynx (ΔD_mean_[Gy], ΔV_50Gy_[%]), mandible (ΔD_max_[Gy]), CTV high (ΔD_1%_[Gy], ΔD_95%_[Gy]), CTV intermediate (ΔD_1%_[Gy], ΔD_95%_[Gy]), and CTV low (ΔD_1%_[Gy], ΔD_95%_[Gy]). In both the comparison and ablation studies, we compared model performance by computing performance metrics on the validation set data using the model saved during training. The model chosen to predict dose for the final test set was the model that performed the best in the ablation study. Boxplots of metrics were created to show the final selected model’s test set performance. A median dose volume histogram was also computed using the final selected model’s performance on the test set cohort.

## Results

A total of 62 patients were identified for model training and evaluation. The training set consisted of 41 patients, the validation set 10 patients, and the test set 11 patients.

A summary of the comparison study results can be found in Table 1. The results in the table indicate that our model had the best performance compared to the two comparison models. This is indicated by our model’s superior performance in all imaging metrics evaluated (MAE, SSIM, and PSNR) and most clinical metrics, such as the spinal cord ΔD_max_[Gy] and CTV High ΔD_95%_[Gy].

**Table 1:**
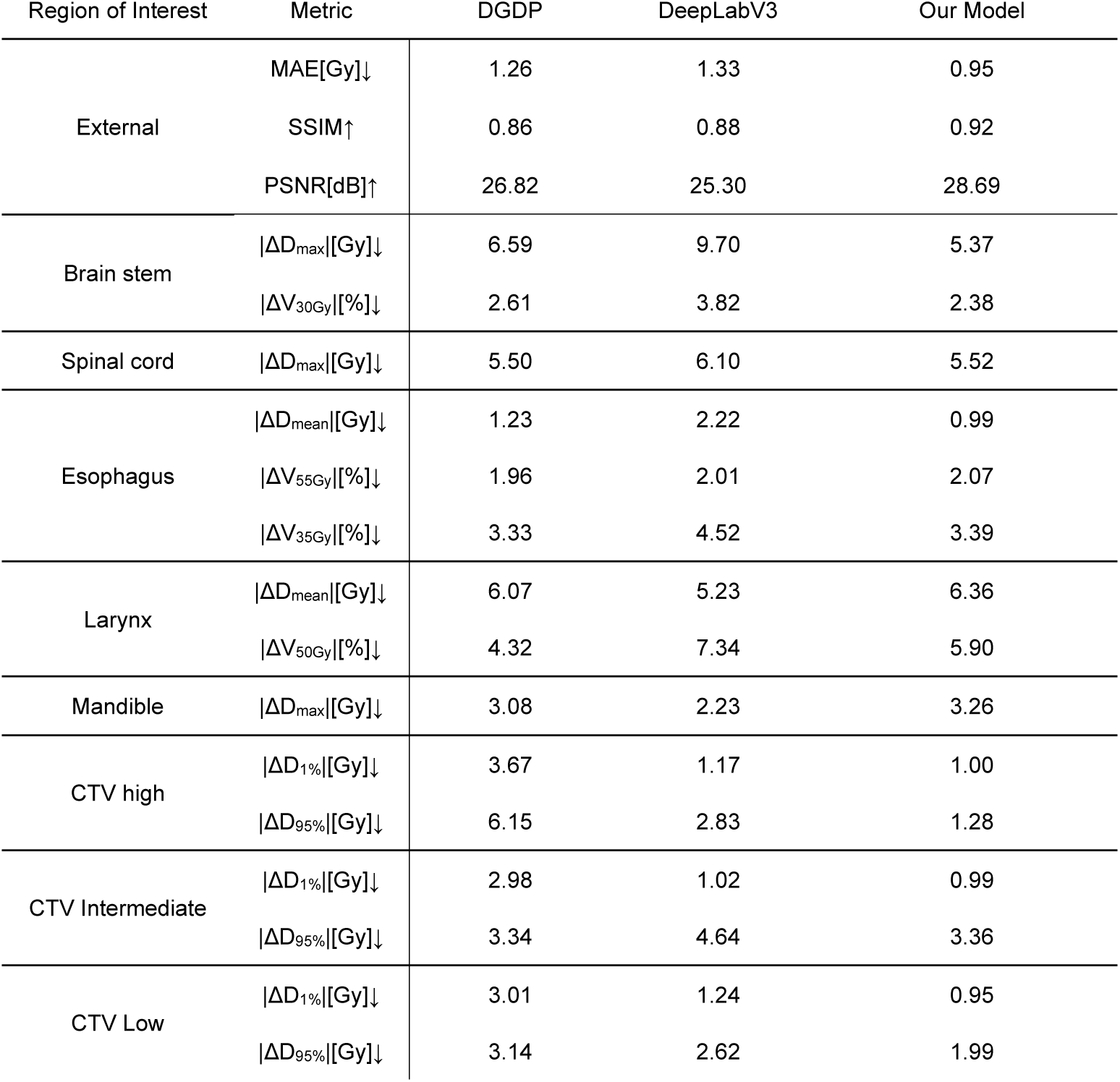
Summary of comparison study results.

A summary of the ablation study results can be found in Table 2. The results in the table indicate that the best performing model was the model incorporating all tested aspects including generated field masks, SDMs, ROIs, and four field predictions. The model with all components had the best performance on all imaging metrics and most of the clinical metrics.

**Table 2:**
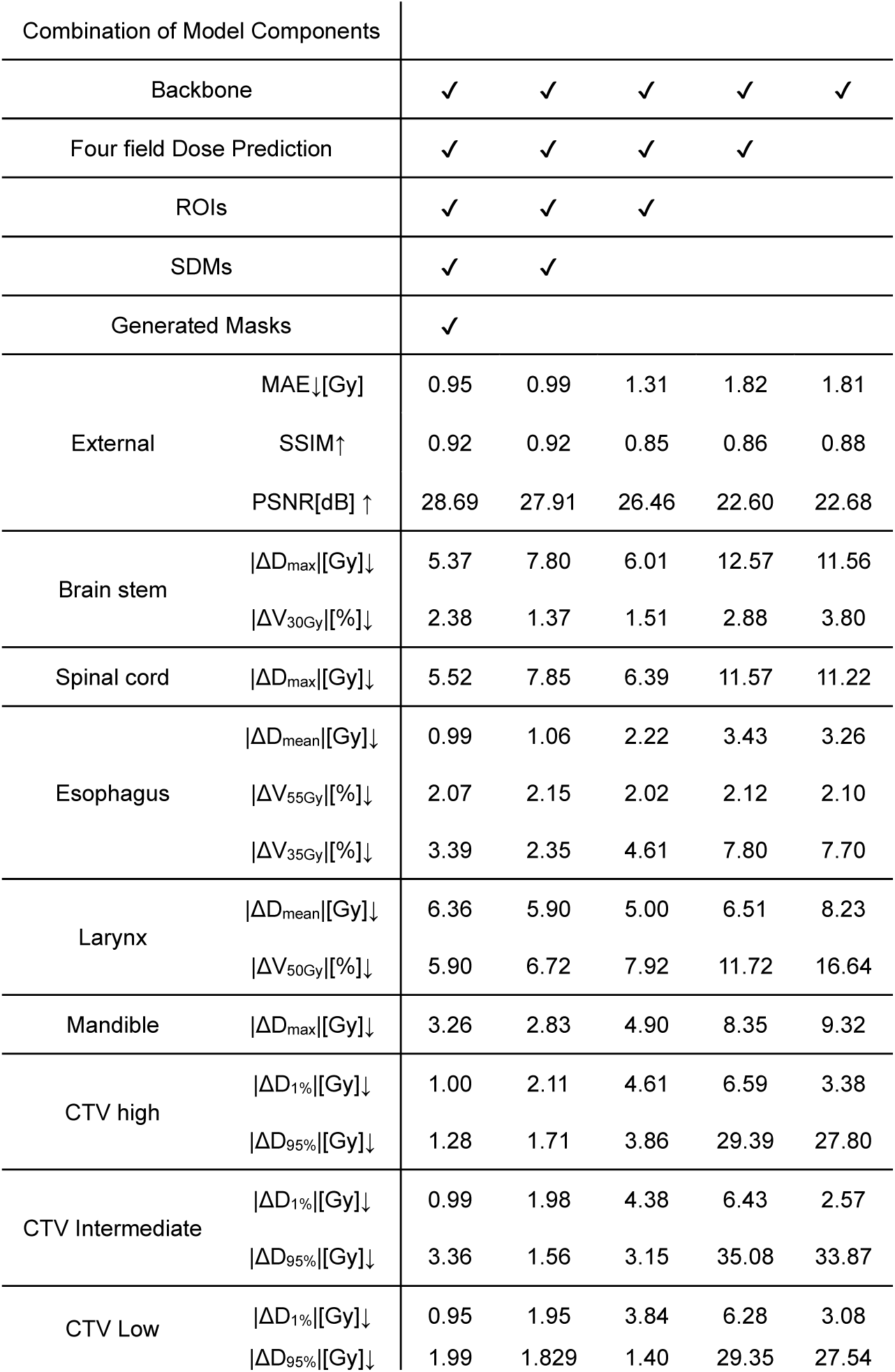
Summary of ablation study results.

| Combination of Model Components |  |  |  |  |  |  |
| --- | --- | --- | --- | --- | --- | --- |
| Backbone |  | ✓ | ✓ | ✓ | ✓ | ✓ |
| Four field Dose Prediction |  | ✓ | ✓ | ✓ | ✓ |  |
| ROIs |  | ✓ | ✓ | ✓ |  |  |
| SDMs |  | ✓ | ✓ |  |  |  |
| Generated Masks |  | ✓ |  |  |  |  |
| External | MAE↓[Gy] | 0.95 | 0.99 | 1.31 | 1.82 | 1.81 |
|  | SSIM↑ | 0.92 | 0.92 | 0.85 | 0.86 | 0.88 |
|  | PSNR[dB] ↑ | 28.69 | 27.91 | 26.46 | 22.60 | 22.68 |
| Brain stem | $\vert \Delta D_{\max}\vert $[Gy]↓ | 5.37 | 7.80 | 6.01 | 12.57 | 11.56 |
| | $\vert \Delta V_{30\text{Gy}}\vert $[%]↓ | 2.38 | 1.37 | 1.51 | 2.88 | 3.80 |
| Spinal cord | $\vert \Delta D_{\max}\vert $[Gy]↓ | 5.52 | 7.85 | 6.39 | 11.57 | 11.22 |
| Esophagus | $\vert \Delta D_{\text{mean}}\vert $[Gy]↓ | 0.99 | 1.06 | 2.22 | 3.43 | 3.26 |
| | $\vert \Delta V_{55\text{Gy}}\vert $[%]↓ | 2.07 | 2.15 | 2.02 | 2.12 | 2.10 |
| | $\vert \Delta V_{35\text{Gy}}\vert $[%]↓ | 3.39 | 2.35 | 4.61 | 7.80 | 7.70 |
| Larynx | $\vert \Delta D_{\text{mean}}\vert $[Gy]↓ | 6.36 | 5.90 | 5.00 | 6.51 | 8.23 |
| | $\vert \Delta V_{50\text{Gy}}\vert $[%]↓ | 5.90 | 6.72 | 7.92 | 11.72 | 16.64 |
| Mandible | $\vert \Delta D_{\max}\vert $[Gy]↓ | 3.26 | 2.83 | 4.90 | 8.35 | 9.32 |
| CTV high | $\vert \Delta D_{1\%}\vert $[Gy]↓ | 1.00 | 2.11 | 4.61 | 6.59 | 3.38 |
| | $\vert \Delta D_{95\%}\vert $[Gy]↓ | 1.28 | 1.71 | 3.86 | 29.39 | 27.80 |
| CTV Intermediate | $\vert \Delta D_{1\%}\vert $[Gy]↓ | 0.99 | 1.98 | 4.38 | 6.43 | 2.57 |
| | $\vert \Delta D_{95\%}\vert $[Gy]↓ | 3.36 | 1.56 | 3.15 | 35.08 | 33.87 |
| CTV Low | $\vert \Delta D_{1\%}\vert $[Gy]↓ | 0.95 | 1.95 | 3.84 | 6.28 | 3.08 |
| | $\vert \Delta D_{95\%}\vert $[Gy]↓ | 1.99 | 1.829 | 1.40 | 29.35 | 27.54 |

The best performing model’s test set imaging metrics performance was the following (mean±SD): MAE 1.02±0.26 Gy, SSIM 0.91±0.02 and PSNR 28.50±2.32 dB. Figure 3 shows boxplots of the best performing model’s test set clinical metrics. Table 3 contains the test set metrics for each of the individual fields. The median dose volume histogram for the final dose prediction model and the various organs at risk is shown in Figure 4. Figure 5 shows dose prediction examples for both field-level and total dose within the test set using the final chosen model.

**Figure 3:**
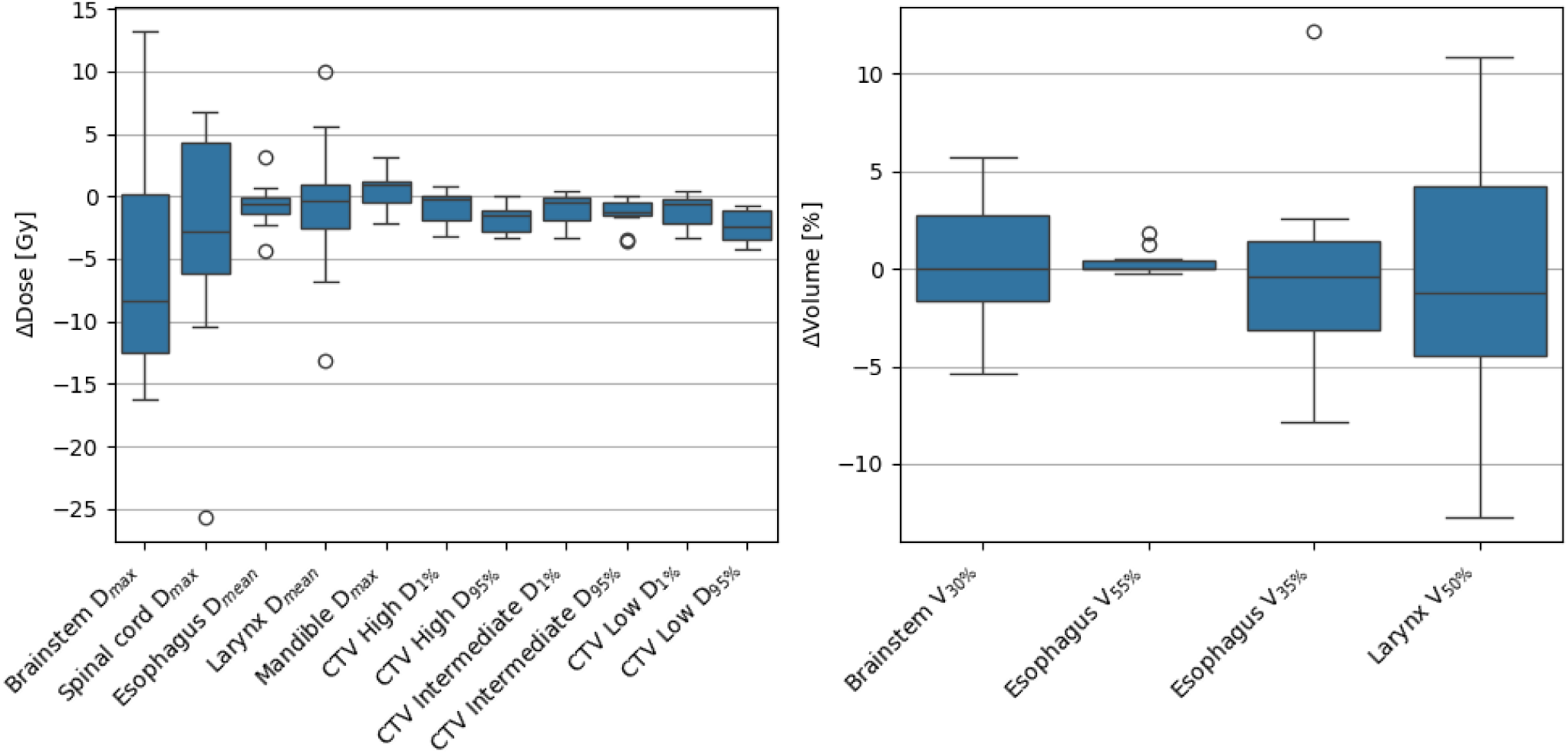
Boxplots of test set prediction metrics using the final field geometry aware model. The left plot shows dose-based DVH metrics. The right plot shows volume-based DVH metrics. The difference for each metric is computed as ground truth – prediction.

**Figure 4:**
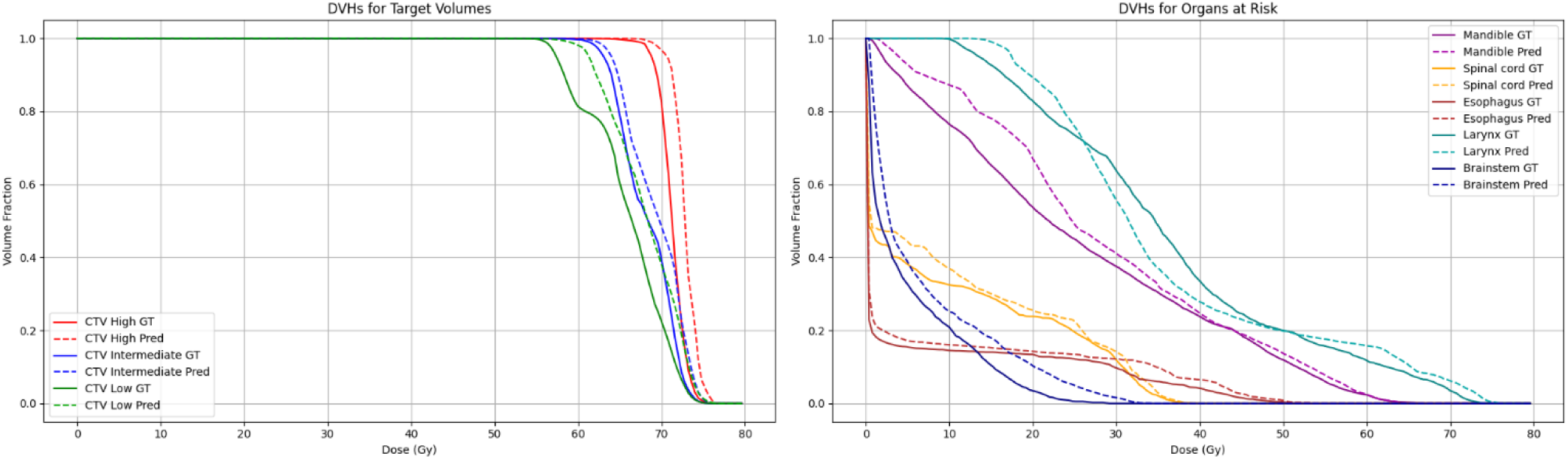
Summary of final chosen model cumulative DVH curves for targets and organs at risk. The left plot shows targets (CTVs) and the right plot shows organs at risk. Each point corresponds to the median value of all patient DVHs for that point on the DVH curve. Solid lines correspond to ground truth (GT) and dotted lines correspond to predictions (pred). The line corresponds to the median cumulative DVH component. Cases with blank ROI information were excluded when computing the graph.

**Figure 5:**
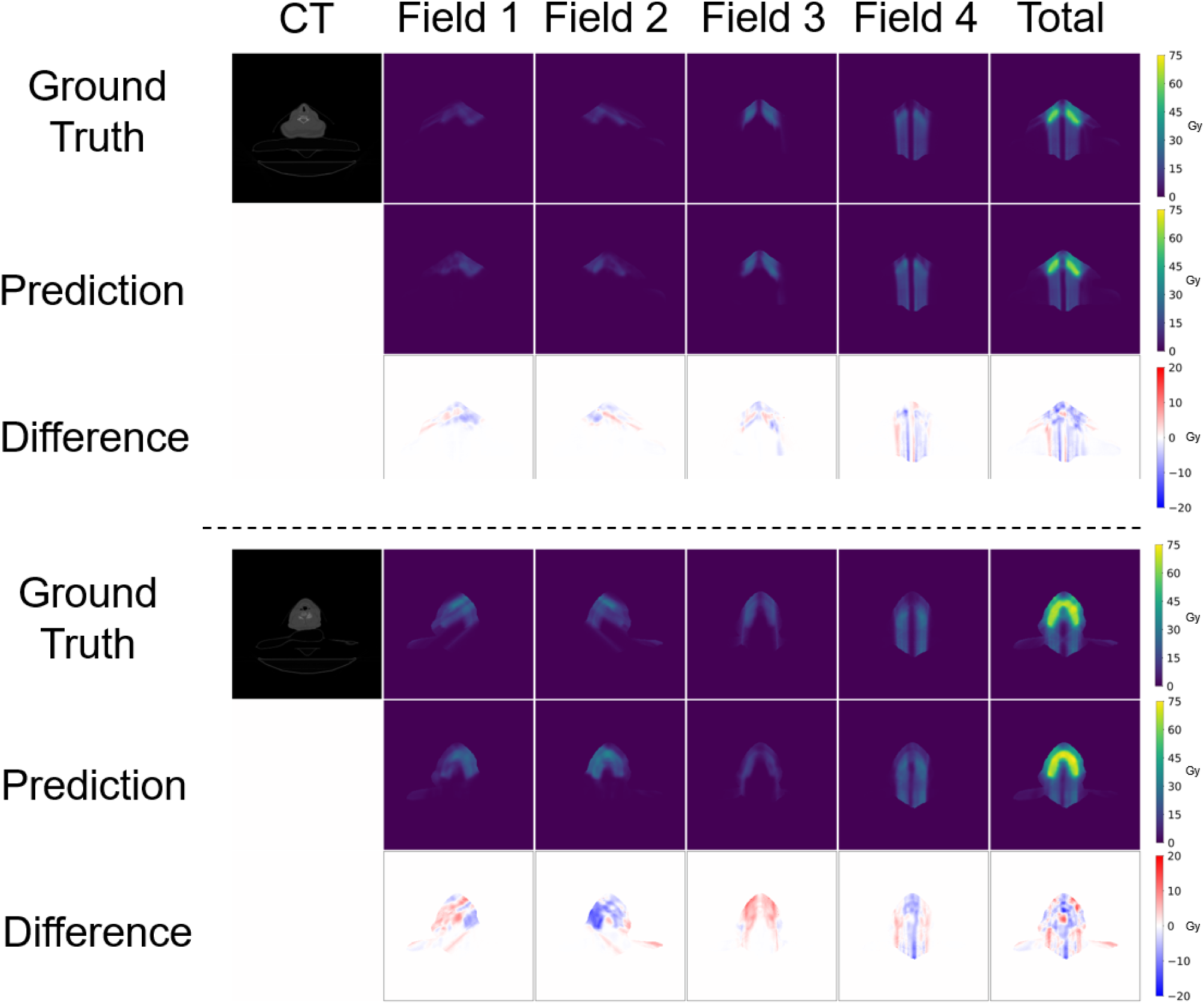
Examples of four field dose predictions using the final dose prediction model. Examples for two different cases are shown. The total predicted dose is computed by summing the four field doses together.

**Table 3:**
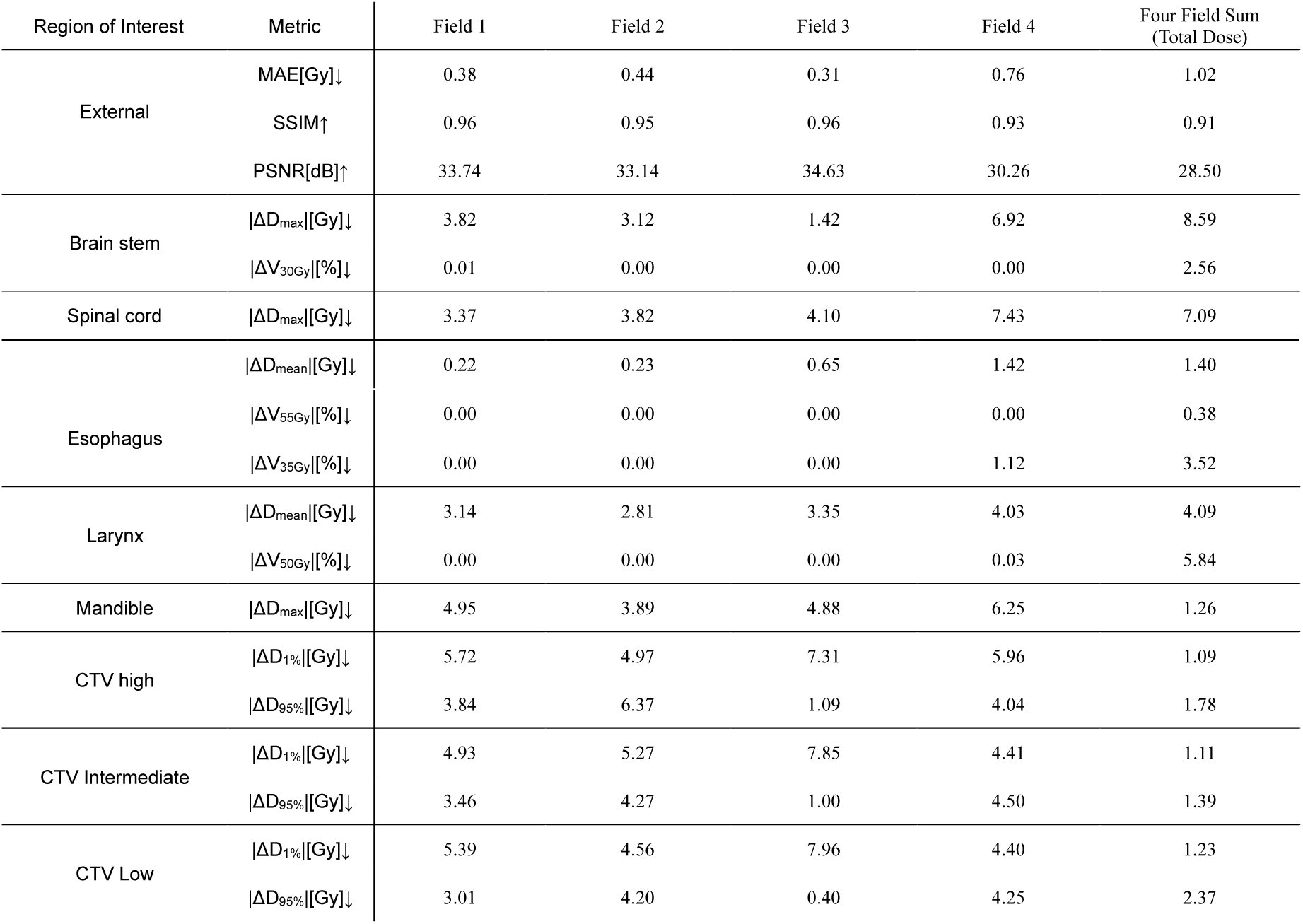
Summary of study results of the data on the test set.

## Discussion

Deep learning techniques have become a popular choice in recent years for head and neck cancer radiation therapy dose prediction tasks. In comparison to photon HNC treatment plans, most proton plans consist of a series of treatment fields rather than treatment arcs. For proton deep learning-based dose prediction, this field geometry information is frequently neglected in both the model input and in the prediction of proton field outputs. Without incorporating this information, the optimal treatment plan that is clinically deliverable may not be generated. In addition, to date, no published HNC proton dose prediction model has yet to incorporate image-based transformers, despite image-based transformers proving successful in radiation therapy dose prediction for other treatment sites^29 30^.

In this study, we developed a head and neck cancer proton radiation therapy dose prediction model that incorporates field geometry information in both model input and output. For model input, field geometry is included by providing proton field masks and field SDMs. For model output, field geometry is considered by training the model to output the dose for each radiation treatment field. Our model can be used to generate field contributions to a patient’s treatment plan or the total dose by summing all treatment fields together. In addition, the model incorporates a cross-attention transformer that has yet to be used for per proton field head and neck cancer dose prediction.

The input to our model uses field masks for each of the four proton fields. These beam masks are not created from the ground truth fields, which would not be available during model inference. Instead, the beam masks are created using the gantry angle and couch angle in a procedure shown in Figure S1. For conventionally planned treatments, the gantry angle and treatment couch angle are typically chosen by the dosimetrist. Our model can be used to plan treatments by either using dosimetrist chosen angles or by using the mean gantry rotation and treatment couch rotation angles based on previous plans. A rapid set of plans can be generated by varying treatment couch and gantry angles. This rapid plan generation may accelerate choosing the final angles as the dose prediction output is quicker compared to traditional planning methods.

Table 1 shows a summary of the evaluation metrics between the two comparison models and our developed model. Our developed proton dose prediction model showed superior performance for all three of the imaging metrics (MAE, SSIM, and PSNR) evaluated. For clinical metrics, our developed model outperformed the other models for most metrics. In particular, for most CTV metrics, our model outperformed the other models.

For our model, we chose to use 2D slices for model input rather than full 3D images. This choice was made for several reasons. First, a 2D model allows for a more flexible input size along the axial axis. Many CTs do not have similar axial scan lengths. By using a model with 2D input, various scan lengths can be used without the need to resample or crop the image. In addition, 2D models are smaller than full 3D models which allows for greater flexibility when using the models clinically. A potential disadvantage to using a 2D model is that each slice by itself cannot use the full 3D imaging context of the slice. The SDMs included in the final model should help to overcome this issue. The SDMs provide for each slice 3D spatial information that would not be available to the model otherwise.

Our model showed performance improvements as measured by MAE with the addition of each model component. A large performance improvement occurred when signed distance maps were added to the model. This performance improvement may be attributed to the signed distance maps providing the model three-dimensional information that is absent from the two-dimensional images alone. The final model with all components included had the best final model performance on all three imaging metrics (MAE, SSIM, PSNR) and the best performance on most clinical metrics.

In the median cumulative DVH, the model had strong performance compared to the ground truth test set data. The largest discrepancy between the predicted and ground truth DVH was for the lower dose (10-20 Gy) region of the mandible. In this region, the model predicted slightly hotter dose distributions compared to the ground truth. This may have occurred because of slight differences between the training and test sets. Although care was taken to match the two sets of dose distributions, the training set may have had slightly warmer plans overall.

A potential limitation of this study was the use of a single institution dataset for model training and evaluation. Although this may limit overall generalizability, this approach is beneficial for our institution’s potential clinical use of the model. Furthermore, another potential limitation is that our model produces dose predictions for a single head and neck cancer subsite, bilateral oropharynx cancer, using a single dose protocol. In future work, we hope to expand our model to predict proton field dose for additional HNC subsites and dose protocols.

## Conclusion

In this work, we created a head and neck cancer proton dose prediction model that can predict per field radiation dose. Our model can be used to evaluate field-level contributions to a patient’s treatment or the entire plan. To construct our model, we incorporated radiation field geometry information to help construct plans that may be more clinically relevant compared to single total field predictions alone. Our model demonstrated strong dose prediction performance on a variety of imaging and clinical metrics. The final model has the potential to be used clinically to improve the quality and speed of head and neck cancer proton radiation therapy plan development.

## Author contributions (CRediT Statement)

In accordance with the Contributor Roles Taxonomy (CRediT, https://credit.niso.org/), the contributing authors have designated responsibilities and individual author attribution. The corresponding author(s) assume(s) responsibility for role assignment, and all contributors have been given the opportunity to review and confirm assigned roles. <u>Brandon Reber</u>: Conceptualization, Methodology, Software, Writing – Original Draft <u>Satomi Shiraishi</u>: Supervision, Conceptualization, Writing – Review and Editing <u>Andrew Foong</u>: Methodology, Writing – Review and Editing, <u>David Routman</u>: Data Curation, Writing – Review and Editing, <u>Jing Qing</u>: Supervision, Conceptualization, Methodology, Writing – Review and Editing

## Institutional Review Board Statement

The institutional review board 22-005097 determined that this work was exempt from the IRB review board approval requirement (45 CFR 46.104d, category 4), on 6/2/2022.

## Data availability Statement

Research data is subject to patient data privacy laws that protect data from being published externally.

## Supporting Documentation

**Figure S1:**
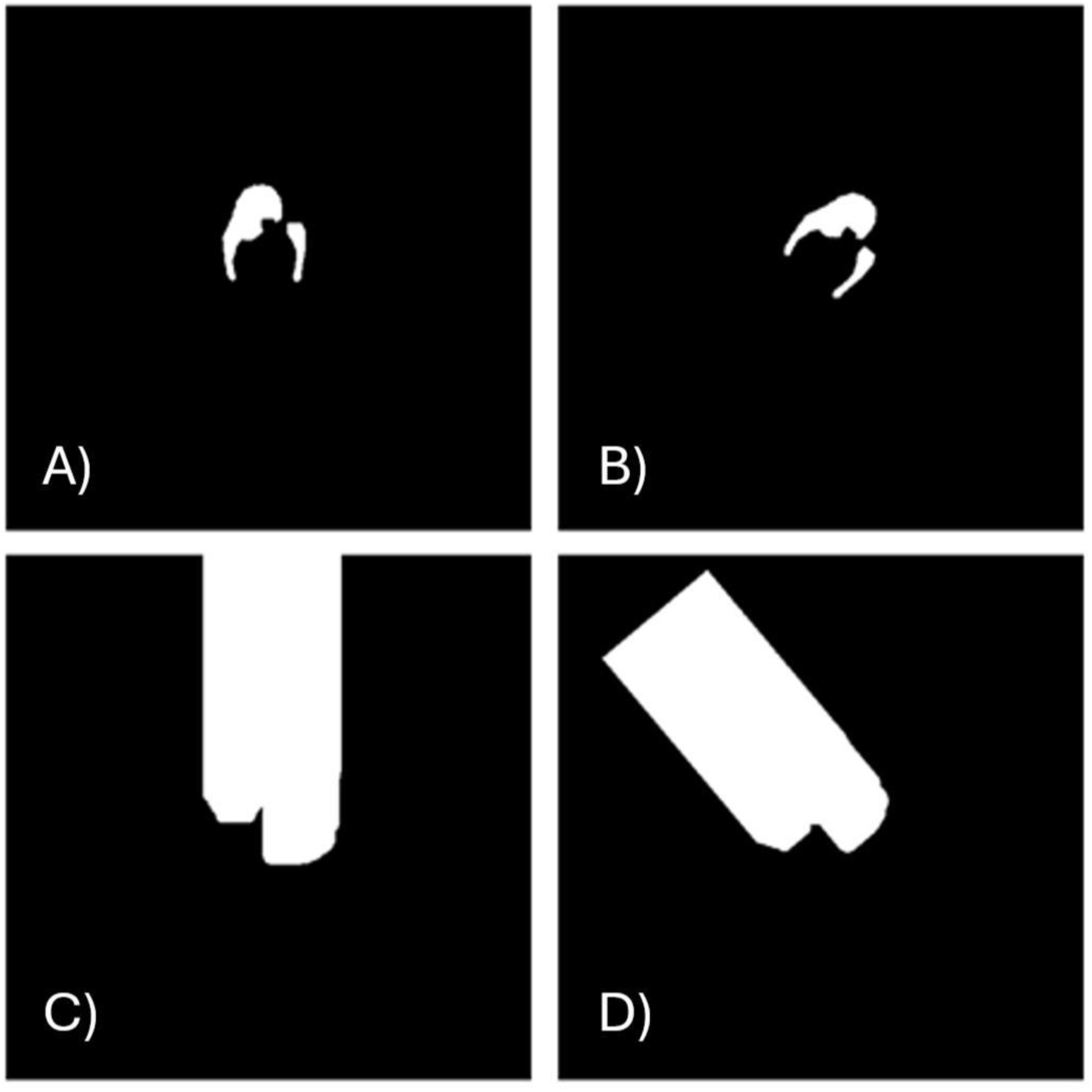
Beam mask generation. Each of the four sections shows an axial slice of the beam mask at a different step in the beam generation process. To generate the beam masks, the treatment isocenter, gantry angle, and couch rotation angle are collected. (A) A combined mask that is the union of CTV High, CTV Intermediate, and CTV Low masks was collected. The center of the image array was shifted to the treatment isocenter. (B) The array is then rotated by the gantry angle followed by the couch rotation angle. The max and min x and z indices containing mask voxels were identified. Additionally, the largest nonzero y index (bottom y voxel) was collected. (C) Voxels starting at the bottom y voxel are filled upward bounded by the x and z indices found in the previous step. (D) The array is rotated backward by the couch rotation angle and gantry angle, and the image isocenter shifted back to its original location.

**Figure S2.**
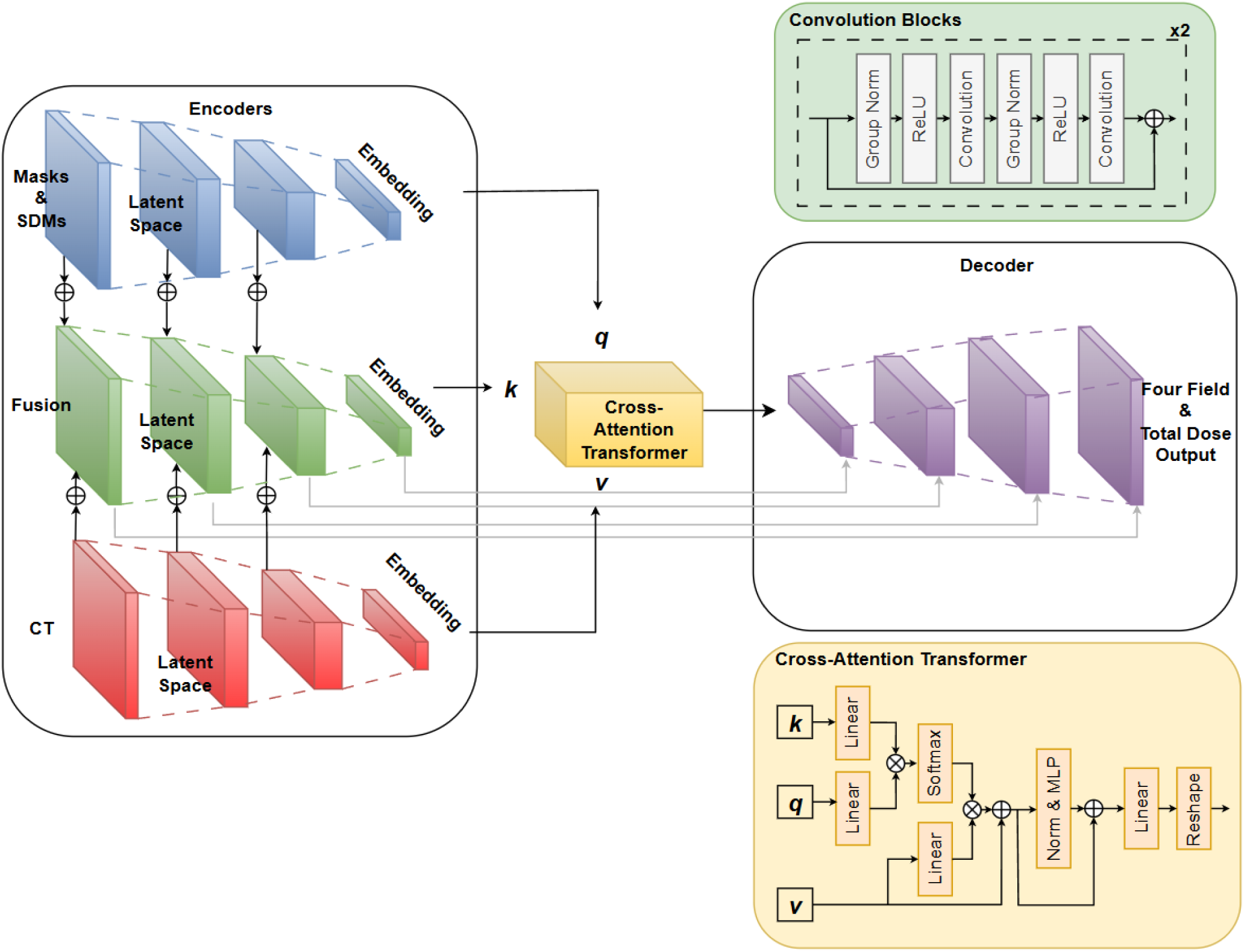
Deep learning architecture with additional architecture details. The green top right diagram describes the residual blocks used at each feature resolution for both the encoder and the decoder branches. The bottom right diagram shows the cross-attention transformer components. Here, MLP stands for multilayer perceptron.

## Additional architecture construction details

The input to the deep learning model with all model components was one CT channel and 28 channels for oar masks, OAR signed distance maps, generated beam masks, and generated beam signed distance maps. For each encoder input, a 1x1 convolution creates 128 channels for each of the three encoder branches. The encoder and decoder branches each consist of four stages. The number of channels at each stage is doubled for the encoder branches or halved for each stage of the decoder branch. Each stage consists of two residual blocks. Each residual block consists of a group normalization layer, a ReLU layer, a 3x3 convolution layer, a ReLU layer, and a 3x3 convolution layer. Skip connections sum the output of each residual block to the block input. For the encoder branches, after the two residual blocks, a 3x3 convolution layer with a stride of 2 downsamples the features by half. For the decoder branch, after the second residual block, nearest neighbor interpolation is completed that doubles the resolution. Skip connections sum decoder stage input to the corresponding fused feature encoder branch stage output. At the end of the model, a 1x1 convolution layer produces four outputs corresponding to each of the field maps.

The bottleneck of the model uses a cross-attention transformer. A summary of the transformer is in the lower right portion of Figure S2. The cross-attention transformer uses the fused feature encoder features as keys, the mask encoder branch features as queries, and the CT encoder branch features as values. Each of the branches first undergoes patch embedding using a patch size of 8x8 and dimension 1024. Position embedding is then learned during training and summed to the patch embeddings. The tokens then undergo LayerNorm followed by a linear layer. The key and query tokens are then multiplied with each other elementwise followed by a softmax activation function. Next, the output of the softmax activation function is multiplied elementwise to the value tokens followed by summation with the value tokens. The output of this process is used as input to a multilayer perceptron (MLP). The MLP consists of a linear layer with 2048 output channels, a GELU layer, a linear layer with 1024 output channels, and a GELU layer. After the MLP, LayerNorm is used followed by summing with the initial MLP input. Next, a linear layer is used followed by a reshaping of the transformer output to the original encoder feature dimensions.

## Distance guided dose prediction (DGDP) architecture and training details

The best performing model in the distance-guided dose prediction paper was the model using the transformed signed distance boundary maps (TSDBMs). The DGDP comparison model uses TSDBM inputs of the brainstem, esophagus, larynx, left parotid, right parotid, external, CTV high, CTV intermediate, CTV low, and spinal cord. Matching the training parameters used in the original paper, the L1 loss function was used with a batch size of 10, initial learning rate of 1e-4, 200 epochs, and Adam optimizer. The learning rate is held constant for the first 100 epochs before decaying linearly to 1e-7. The validation set was used to select the best performing model during training using the L1 loss function between the predicted dose and ground truth dose.

## DeepLabV3 architecture and training details

The input to the DeepLabV3 architecture was the treatment planning CT and nine OAR masks. The nine OAR masks were the masks of the brainstem, esophagus, larynx, left parotid, right parotid, mandible, spinal cord, external, and CTV mask. The base of the model uses the built-in PyTorch (version 2.1.0+cu121) function deeplabv3_resnet50 with default weights. The first layer that takes in the input was changed to accept the concatenated CT and masks. The final classifier layer was changed to a convolution layer using a 1x1 kernel. The mean squared error loss function was used. The model was trained for 500 epochs in total. A step learning rate scheduler was used with the learning rate decreased by a factor of 10 every 125 epochs. Models were trained using four Nvidia A100 GPUs with a batch size of 64 split between the four GPUs. The validation set was used to select the best performing model during training using the mean squared error between the predicted dose and ground truth dose.

